# Reproductive Experiences and Cardiovascular Disease Care in Pregnancy Capable and Post-Menopausal Individuals: Insights from the American Heart Association Research Goes Red Registry

**DOI:** 10.1101/2023.03.14.23287279

**Authors:** Shiavax J. Rao, Yaa A. Kwapong, Ellen Boakye, Pratheek Mallya, Juan Zhao, William Akel, Haoyun Hong, Shen Li, Chigolum P. Oyeka, Faith Elise Metlock, Pamela Ouyang, Roger S. Blumenthal, Khurram Nasir, Abha Khandelwal, Claire Kinzy, Laxmi S. Mehta, Veronique L. Roger, Jennifer L. Hall, Garima Sharma

## Abstract

**Background:** Information on reproductive experiences and awareness of adverse pregnancy outcomes (APOs) and cardiovascular disease (CVD) risk among pregnancy-capable and post-menopausal individuals has not been well described. We sought to evaluate preconception health and APO awareness in a large population-based registry.

**Methods:** Data from the Fertility and Pregnancy Survey of the American Heart Association Research Goes Red Registry (AHA-RGR) were used. Responses to questions pertaining to prenatal health care experiences, postpartum health, and awareness of the association of APOs with CVD risk were used. We summarized responses using proportions for the overall sample and by stratifications, and we tested differences using the Chi-squared test.

**Results:** Of 4,651individuals in the AHA-RGR registry, 3,176 were of reproductive age, and 1,475 were postmenopausal. Among postmenopausal individuals, 37% were unaware that APOs were associated with long-term CVD risk. This varied by different racial/ethnic groups (non-Hispanic White: 38%, non-Hispanic Black: 29%, Asian: 18%, Hispanic: 41%, Other: 46%; *P* = 0.03). Fifty-nine percent of the participants were not educated regarding the association of APOs with long-term CVD risk by their providers. Thirty percent of the participants reported that their providers did not assess pregnancy history during current visits; this varied by race-ethnicity (*P* = 0.02), income (*P* = 0.01), and access to care (*P* = 0.02). Only 37.1% of the respondents were aware that CVD was the leading cause of maternal mortality.

**Conclusions:** Considerable knowledge gaps exist in the association of APOs with CVD risk, with disparities by race/ethnicity, and most patients are not educated on this association by their health care professionals. There is an urgent and ongoing need for more education on APOs and CVD risk, to improve the health-care experiences and postpartum health outcomes of pregnant individuals.

## Introduction

Cardiovascular disease (CVD) accounts for over 400,000 deaths annually in the United States (US), making it a leading cause of morbidity and mortality in women.^1^ Despite these alarming statistics, a national survey of women’s CVD awareness conducted by the American Heart Association (AHA) in 2019 reported a significant decline in knowledge and awareness regarding heart disease as a leading cause of death over the last decade,^2^ most noticeable among younger women and certain racial/ethnic groups. Approximately 20% of coronary heart disease events may occur in the absence of conventional risk factors,^3^ and current stratification tools might be inadequate for cardiovascular (CV) risk assessment in younger women.^4^ To address this disparity, sex-specific CV risk factors, such as adverse pregnancy outcomes (APOs), can be utilized for CVD risk assessment in this population.^5,6^

Pregnancy is associated with a physiological challenge to the CV system with increased circulating volume, hyperlipidemia, insulin resistance, and an increase in inflammatory and clotting factors.^7^ Approximately 10-20% of women experience APOs – including pre-term birth, preeclampsia, and intrauterine growth restriction – which are common interrelated disorders caused by placental dysfunction and maternal vascular abnormalities including endothelial activation, inflammation, and vasospasm.^8,9^ In addition, pregnancy-capable individuals are at an increased risk for developing future CVD, as APOs are associated with an increased risk of development of hypertension, left ventricular hypertrophy/dysfunction, vascular dysfunction, and renal dysfunction.^8,9^

The awareness among physicians and patients of the association between APOs and future CVD is not well-described. Responses from a recent survey completed by physicians in several specialties revealed that only half of internists and family physicians acknowledged awareness of this association,^8^ with the majority of physicians in internal medicine, family medicine and cardiology demonstrating a lack of knowledge regarding how often to appropriately screen for CVD risk factors associated with APOs.^8^ In our study, we examined responses of pregnancy-capable individuals to questions in the Fertility and Pregnancy Survey from the AHA Research Goes Red (RGR) Registry^10^ to (1) understand experiences of preconception and pregnancy care, (2) evaluate post-conception health including postpartum depression (PPD), (3) understand awareness of the association between APOs and long-term CVD risk, (4) understand provider counseling of patients on the association between APOs and long-term CVD risk, and (5) evaluate whether healthcare providers include pregnancy history in taking medical history on routine visits.

## Methods

### Study design and population sample

In this cross-sectional study, we utilized data from participants enrolled in the AHA RGR Registry, which was launched in collaboration with Verily’s Project Baseline.^10^ The RGR is a longitudinal dynamic registry with several targeted health surveys available to consenting adults aged ≥ 18 years in the US. Project Baseline is an effort to expedite evidence generation in clinical research. Established in 2019, RGR is a novel online platform designed to be participant-centric, customizable, and scalable. Individuals registered for RGR from the Project Baseline website (projectbaseline.com). The website and Clinical Studies Platform were provided by Verily. The study utilized data from 4,651 participants who had responded to the Fertility and Pregnancy Survey **(appendix 1)** as of September 2022. There were two subgroups of participants: individuals of the reproductive age group (18 – 44 years, n = 3,176) and individuals who were no longer of childbearing age (≥45 years, n = 1,475).

Reproductive status was assessed with the question “What best describes you now?” Reproductive age group included participants who were currently pregnant, postpartum (pregnant within the last 12 months), actively trying to get pregnant, or considering pregnancy. Participants who answered “I have been pregnant and/or had children but am no longer of childbearing age/status” were considered postmenopausal.

This study was approved by the Western Institutional Review Board (#101143) – The Project Baseline Community Study. We followed the Strengthening the Reporting of Observational Studies in Epidemiology guidelines in reporting our findings.^11^

### Variables assessed

Awareness of Adverse Pregnancy Outcomes and Long-term Cardiovascular Risk was assessed in postmenopausal individuals, specifically individuals who had been pregnant and/or had children but were no longer of childbearing age/status. Preconception Health and Prenatal Care was assessed among individuals of reproductive age. Postpartum health, specifically postpartum depression (PPD) was assessed in postpartum individuals. Sociodemographic characteristics assessed included age, race/ethnicity, educational status, annual household income, and proximity to healthcare.

### Statistical analysis

Sociodemographic characteristics of all study participants were summarized by proportions. Responses of postmenopausal individuals on awareness of adverse pregnancy outcomes and cardiovascular risk and provider counseling on these associations were also summarized using proportions for the overall samples, and stratified by race/ethnicity, education, income, and proximity to healthcare. Differences among these subgroups were evaluated using the Chi-square test. Responses for all participants of reproductive age regarding preconception health and prenatal care were summarized using proportions. Responses of postpartum participants on PPD screening were summarized using proportions for the overall samples and stratified by race-ethnicity, education, income, and proximity to healthcare. We used fisher’s exact test to examine the differences in PPD screening for different subgroups stratified by race-ethnicity, education, income, and proximity to healthcare. *P*-value<0.05 indicated statistically significant differences. Python (version 3.1.7) was used to derive the data and for tabulated analysis. Statistical analysis was performed using the open-source software R (R Foundation for Statistical Computing, Vienna, Austria). The American Heart Association Precision Medicine Platform (https://precision.heart.org) was used for data analysis.

## Results

### Sociodemographic Characteristics of all Participants

Of the 4,651 participants (mean age 42.92 ± 12.06 years), 3,176 were of reproductive age and 1,475 were postmenopausal. Most participants identified as non-Hispanic White (75.7%) or Black (9.8%). Majority of respondents were aged 35 – 39 years (17.0%), had an advanced degree (36.3%), income of $50,000 – $99,999 (29.2%) and were residents of a medium-sized city with access to more than one hospital and healthcare provider within 15 miles of their homes (43.0%) **(Table 1)**.

**Table 1:** Sociodemographic Characteristics of all Study Participants.

|  | N | % |
| --- | --- | --- |
| **Gender at birth** |  |  |
| Women | 4651 | 100 |
| Men | 0 | 0 |
| **Age, years** |  |  |
| 18-24 | 203 | 4.4 |
| 25-29 | 343 | 7.4 |
| 30-34 | 690 | 14.8 |
| 35-39 | 789 | 17.0 |
| 40-44 | 627 | 13.5 |
| 45-49 | 652 | 14.0 |
| 50-54 | 609 | 13.1 |
| 55-59 | 281 | 6.0 |
| 60-64 | 180 | 3.9 |
| ≥65 | 277 | 6.0 |
| **Race-ethnicity** |  |  |
| Non-Hispanic White | 3531 | 75.9 |
| Non-Hispanic Black | 455 | 9.8 |
| Asian | 145 | 3.1 |
| Hispanic | 341 | 7.3 |
| Others (American Indian/Alaskan native, Native Hawaiian or Pacific Islander, Multiracial, others) | 179 | 3.8 |
| **Highest Education** |  |  |
| Some college or technical school degree or less | 478 | 29.2 |
| College 4+ years (college graduate) | 562 | 34.4 |
| Advanced degree (e.g., Master's, Doctorate) | 594 | 36.3 |
| **Annual Income** |  |  |
| <\$50,000 | 498 | 20.4 |
| \$50,000- \$99,999 | 714 | 29.2 |
| \$100,000- \$149,999 | 536 | 22.0 |
| \$150,000- \$199,999 | 290 | 11.9 |
| \$200,000 or more | 279 | 11.4 |
| Prefer not to answer | 124 | 5.1 |
| **Proximity to healthcare** |  |  |
| Big city with access to many hospitals and healthcare providers within 5 miles of my home | 672 | 25.6 |
| medium-sized city with access to more than one hospital and healthcare provider within 15 miles of my home | 1128 | 43.0 |
| small city with access to at least one hospital and healthcare provider in under 30 minutes | 634 | 24.1 |
| rural area and have to travel more than 30 minutes to access a hospital or healthcare provider | 192 | 7.3 |

### Awareness of APOs and CVD risk among Postmenopausal Individuals

Among postmenopausal individuals, 37% were unaware that APOs were associated with long-term CVD risk, and this varied by different race groups – 430 of 1,127 were non-Hispanic White (NHW: 38%), 49 of 170 were non-Hispanic Black (NHB: 29%), 4 of 18 were Asian (18%), 41 of 100 were Hispanic (41%), and 26 of 56 (46%) individuals were from other ethnicities (*P* = 0.03) **(Table 2, Figure 1A)**. The awareness of the association of APOs with long-term cardiovascular disease did not significantly vary by healthcare access (**supplemental table 1**), educational status (**supplemental table 2)** or income (**supplemental table 3**).

**Table 2:** Awareness of Adverse Pregnancy Outcomes and Cardiovascular Risk among Post-menopausal Individuals by Race/ethnicity.

|  | Total | Non-Hispanic White | Non-Hispanic Black | Asian | Hispanic | Others | p-value |
| --- | --- | --- | --- | --- | --- | --- | --- |
| Pregnancy history taken by provider on current visits |  |  |  |  |  |  |  |
| Yes | 913 (61.9) | 689 (61.1) | 104 (61.1) | 11 (50.0) | 71 (71.0) | 38 (67.8) | 0.02* |
| No | 439 (29.7) | 337 (29.9) | 61 (35.8) | 7 (31.8) | 21 (21.0) | 13 (23.2) |  |
| Don't know | 123 (8.3) | 101 (8.9) | 5 (2.9) | 4 (18.1) | 8 (8.0) | 5 (8.9) |  |
| Awareness of the association of adverse pregnancy outcomes including preeclampsia in general with cardiovascular risk |  |  |  |  |  |  |  |
| Yes | 892 (60.5) | 672 (59.7) | 118 (69.4) | 18 (81.8) | 54 (54.0) | 30 (53.6) | 0.03* |
| No | 550 (37.3) | 430 (38.2) | 49 (28.8) | 4 (18.2) | 41 (41.0) | 26 (46.4) |  |
| Don't know | 33 (2.2) | 25 (2.2) | 3 (1.8) | - | 5 (5.0) | - |  |
| Provider education/counseling on these associations |  |  |  |  |  |  |  |
| Yes | 381 (25.8) | 281 (24.9) | 59 (34.7) | 7 (31.8) | 20 (20.0) | 14 (25.0) | 0.23 |
| No | 863 (58.5) | 666 (59.0) | 90 (52.9) | 13 (59.1) | 62 (62.0) | 32 (57.1) |  |
| Don't know | 231 (15.6) | 180 (15.9) | 21 (12.3) | 2 (9.1) | 18 (18.0) | 10 (17.8) |  |

p-value <0.05 -significant; \* indicates significance.
\*Others- Native Hawaiian and other Pacific Islanders, American Indian and Alaska Natives, mixed race

**Figure 1.**
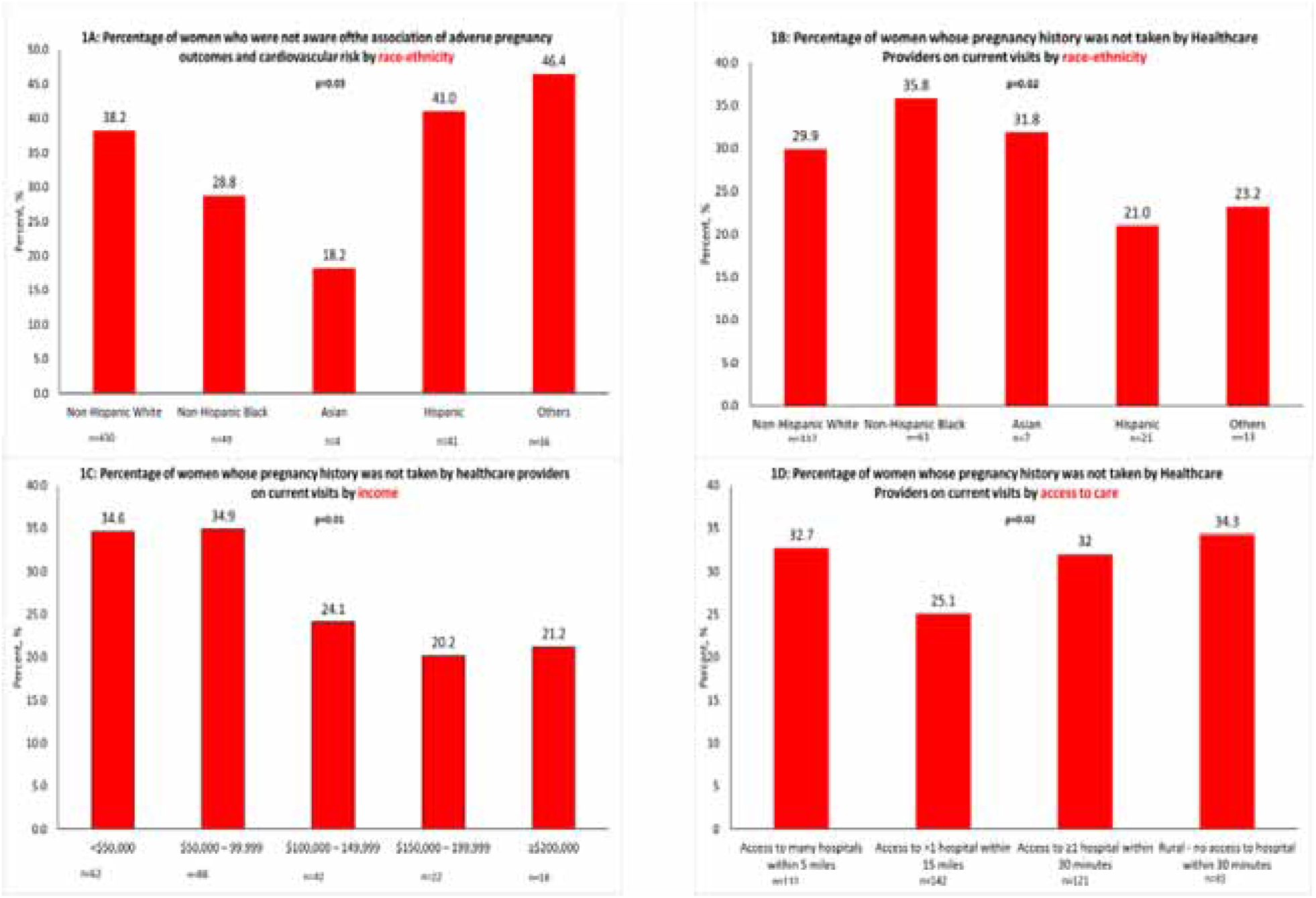
Percentage of postmenopausal women whose pregnancy history were not taken by providers by race/ethnicity, income and access to care

Overall, most (59%) reported that they were not educated on this association by their health care professionals and this proportion varied by educational status: some college or technical school degree or less (54.9%), college graduate (66.8%), advanced degree (61.6%) (*P* <0.05). Additionally, 30% (439 of 1,475) of the postmenopausal individuals reported that their healthcare professionals did not take their pregnancy, reproductive and gynecological history during recent visits, and this proportion significantly varied by race/ethnicity (Figure 1B), income (Figure 1C) and access to care (Figure 1D) (*P* <0.05).

### Preconception Health and Prenatal Care

#### Diagnosis of APO and CVD among Participants of Reproductive Age

Of the total 1,151 participants of reproductive age who responded to related questions on APO and CVD, 10.9% reported a previous diagnosis of preeclampsia and other hypertensive disorders in pregnancy, 4.5% had preterm birth, and 5.7% had gestational diabetes. This was further described by each subcategory (currently pregnant, postpartum, trying to be pregnant, and considering pregnancy) (**Table 3)**.

**Table 3:**
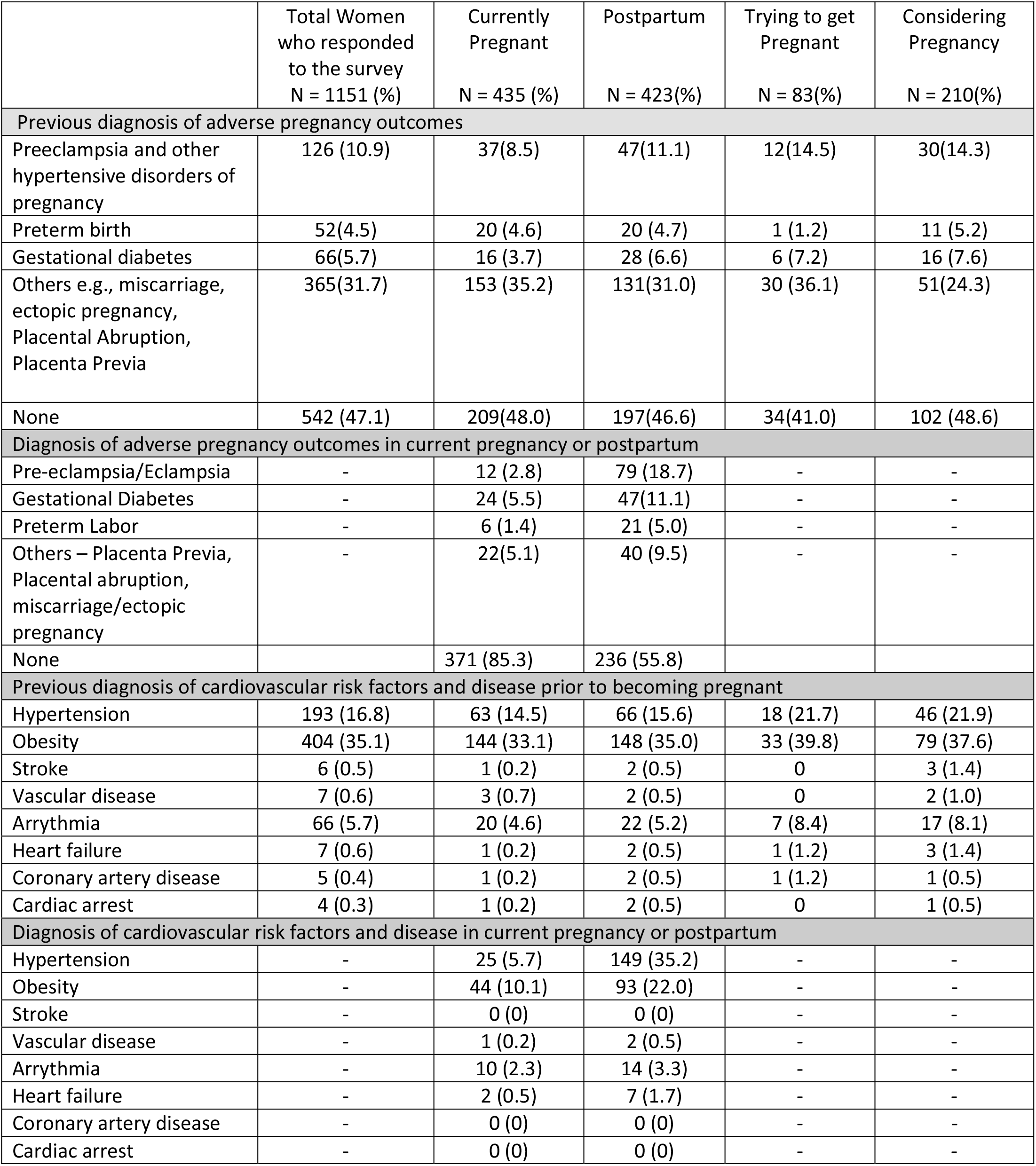
Adverse Pregnancy, Cardiovascular Risk Factors, and Disease among Respondents of Reproductive Age.

|  | Total Women<br>who responded<br>to the survey<br>N = 1151 (%) | Currently<br>Pregnant<br>N = 435 (%) | Postpartum<br>N = 423(%) | Trying to get<br>Pregnant<br>N = 83(%) | Considering<br>Pregnancy<br>N = 210(%) |
| --- | --- | --- | --- | --- | --- |
| **Previous diagnosis of adverse pregnancy outcomes** |  |  |  |  |  |
| Preeclampsia and other<br>hypertensive disorders of<br>pregnancy | 126 (10.9) | 37(8.5) | 47(11.1) | 12(14.5) | 30(14.3) |
| Preterm birth | 52(4.5) | 20 (4.6) | 20 (4.7) | 1 (1.2) | 11 (5.2) |
| Gestational diabetes | 66(5.7) | 16 (3.7) | 28 (6.6) | 6 (7.2) | 16 (7.6) |
| Others e.g., miscarriage,<br>ectopic pregnancy,<br>Placental Abruption,<br>Placenta Previa | 365(31.7) | 153 (35.2) | 131(31.0) | 30 (36.1) | 51(24.3) |
| None | 542 (47.1) | 209(48.0) | 197(46.6) | 34(41.0) | 102 (48.6) |
| **Diagnosis of adverse pregnancy outcomes in current pregnancy or postpartum** |  |  |  |  |  |
| Pre-eclampsia/Eclampsia | - | 12 (2.8) | 79 (18.7) | - | - |
| Gestational Diabetes | - | 24 (5.5) | 47(11.1) | - | - |
| Preterm Labor | - | 6 (1.4) | 21 (5.0) | - | - |
| Others – Placenta Previa,<br>Placental abruption,<br>miscarriage/ectopic<br>pregnancy | - | 22(5.1) | 40 (9.5) | - | - |
| None |  | 371 (85.3) | 236 (55.8) |  |  |
| **Previous diagnosis of cardiovascular risk factors and disease prior to becoming pregnant** |  |  |  |  |  |
| Hypertension | 193 (16.8) | 63 (14.5) | 66 (15.6) | 18 (21.7) | 46 (21.9) |
| Obesity | 404 (35.1) | 144 (33.1) | 148 (35.0) | 33 (39.8) | 79 (37.6) |
| Stroke | 6 (0.5) | 1 (0.2) | 2 (0.5) | 0 | 3 (1.4) |
| Vascular disease | 7 (0.6) | 3 (0.7) | 2 (0.5) | 0 | 2 (1.0) |
| Arrhythmia | 66 (5.7) | 20 (4.6) | 22 (5.2) | 7 (8.4) | 17 (8.1) |
| Heart failure | 7 (0.6) | 1 (0.2) | 2 (0.5) | 1 (1.2) | 3 (1.4) |
| Coronary artery disease | 5 (0.4) | 1 (0.2) | 2 (0.5) | 1 (1.2) | 1 (0.5) |
| Cardiac arrest | 4 (0.3) | 1 (0.2) | 2 (0.5) | 0 | 1 (0.5) |
| **Diagnosis of cardiovascular risk factors and disease in current pregnancy or postpartum** |  |  |  |  |  |
| Hypertension | - | 25 (5.7) | 149 (35.2) | - | - |
| Obesity | - | 44 (10.1) | 93 (22.0) | - | - |
| Stroke | - | 0 (0) | 0 (0) | - | - |
| Vascular disease | - | 1 (0.2) | 2 (0.5) | - | - |
| Arrhythmia | - | 10 (2.3) | 14 (3.3) | - | - |
| Heart failure | - | 2 (0.5) | 7 (1.7) | - | - |
| Coronary artery disease | - | 0 (0) | 0 (0) | - | - |
| Cardiac arrest | - | 0 (0) | 0 (0) | - | - |

Among respondents who were currently pregnant, 2.8% reported a current diagnosis of preeclampsia/eclampsia and 5.5% reported a current diagnosis of gestational diabetes. Among postpartum individuals, the proportion with current diagnoses of preeclampsia/eclampsia, gestational diabetes and preterm birth were 18.7%, 11.1%, and 5.0%, respectively.

Regarding previous diagnosis of CV risk factors prior to becoming pregnant, obesity (35.1%) and hypertension (16.8%) were the most prevalent.

#### Patient Experiences on Preconception and Prenatal Care and Awareness of Leading Cause of maternal Mortality among Respondents of Reproductive Age

Majority of respondents affirmed regular checkup with an obstetrics and gynecology (OBGYN) office (84.0%) or with their primary care provider (56.2%) **(Table 4**). Most of the respondents had health insurance with some/all pregnancy-related coverage (82.3%) and access to prenatal vitamins (96.5%). Approximately a third of the participants (36.2%) affirmed receiving preconception counseling prior to becoming pregnant.

**Table 4:** Patient Experiences on Preconception and Prenatal Care and Awareness of Leading Cause of maternal Mortality among Respondents of Reproductive Age.

|  | Total Women<br>who responded<br>to the survey<br>N = 1151(%) | Currently<br>Pregnant<br>N = 435(%) | Postpartum<br>N = 423(%) | Trying to<br>get<br>Pregnant<br>N = 83(%) | Considering<br>Pregnancy<br>N = 210(%) |
| --- | --- | --- | --- | --- | --- |
| **Patient Experiences of Preconception and Prenatal Care** |  |  |  |  |  |
| Q3: Type of healthcare provider seen within last 12 months |  |  |  |  |  |
| Regular checkup at primary care provider | 647 (56.2) | 237 (54.5) | 215 (50.8) | 59 (71.1) | 136 (64.8) |
| Regular checkup at OB/Gyn's office | 967 (84.0) | 393 (90.3) | 382 (90.3) | 59 (71.1) | 133 (63.3) |
| Regular checkup with nurse/ midwife/doula | - | 78 (17.9) | 119 (28.1) | 4 (4.8) | - |
| Doctor visit for an illness/chronic condition | 465 (40.4) | 159 (36.6) | 158 (37.4) | 39 (47.0) | 109 (51.9) |
| Doctor visit for an injury | 97 (8.4) | 28 (6.4) | 35 (8.3) | 4 (4.8) | 30 (14.3) |
| Doctor visit for family planning | 244 (21.2) | 66 (15.2) | 102 (24.1) | 22 (26.5) | 54 (25.7) |
| Doctor visit for your mental health | 299 (26.0) | 82 (18.9) | 121 (28.6) | 21 (25.3) | 75 (35.7) |
| Dentist/dental hygienist | 737 (64.0) | 285 (65.5) | 265 (62.6) | 55 (66.3) | 132 (62.9) |
| Other | 80 (7.0) | 30 (6.9) | 24 (5.7) | 11 (13.3) | 15 (7.1) |
| Q7. Ever Experienced Bad Obstetric outcome |  |  |  |  |  |
| Miscarriage | 374 (32.5) | 167 (38.4) | 130 (30.7) | 29 (34.9) | 48 (22.9) |
| Ectopic pregnancy | 26 (2.3) | 10 (2.3) | 12 (2.8) | 2 (2.4) | 2 (1.0) |
| Pregnancy termination | 82 (7.1) | 29 (6.7) | 32 (7.6) | 4 (4.8) | 17 (8.1) |
| Still birth | 14 (1.2) | 3 (0.7) | 7 (1.7) | 2 (2.4) | 2 (1.0) |
| Rather not say | 9 (0.8) | 3 (0.7) | 3 (0.7) | 0 (0) | 3 (1.4) |
| Q8. Personal or family history of Infertility |  |  |  |  |  |
| Yes | 334 (29.0) | 119 (27.4) | 116 (27.4) | 31 (37.3) | 68 (32.4) |
| No | 672 (58.4) | 267 (61.4) | 256 (60.5) | 43 (51.8) | 106 (50.5) |
| Not sure | 145 (12.6) | 49 (11.3) | 51 (12.1) | 9 (10.8) | 36 (17.1) |
| Q10. Access to parental leave/health insurance |  |  |  |  |  |
| Paid maternity/paternity leave | 135 (11.7) | 46 (10.6) | 82 (19.4) | 1 (1.2) | 6 (2.9) |
| Unpaid maternity/paternity leave | 118 (10.3) | 35 (8.0) | 80 (18.9) | 2 (2.4) | 1 (0.5) |
| Short term disability | 111 (9.6) | 44 (10.1) | 59 (13.9) | 2 (2.4) | 6 (2.9) |
| None of the above | 145 (12.6) | 25 (5.7) | 67 (15.8) | 13 (15.7) | 40 (19.0) |
| Health insurance with some/all pregnancy related coverage | 947 (82.3) | 397 (91.3) | 329 (77.8) | 67 (80.7) | 154 (73.3) |
| Health insurance without pregnancy-related coverage | 15 (1.3) | 6 (1.4) | 5 (1.2) | 1 (1.2) | 3 (1.4) |
| Not sure | 20 (1.7) | 4 (0.9) | 5 (1.2) | 2 (2.4) | 9 (4.3) |
| Q12: Access to or taking prenatal vitamins |  |  |  |  |  |
| Yes | 1111 (96.5) | 432 (99.3) | 416 (98.3) | 78 (94.0) | 185 (88.1) |
| No | 38 (3.3) | 3 (0.7) | 6 (1.4) | 5 (6.0) | 24 (11.4) |
| I'm not sure | 2 (0.2) | 0 (0) | 1 (0.2) | 0 (0) | 1 (0.5) |
| Q16: Preconception counselling prior to becoming pregnant |  |  |  |  |  |
| Yes | 417 (36.2) | 154 (35.4) | 145 (34.3) | 40 (48.2) | 78 (37.1) |
| No | 681 (59.2) | 268 (61.6) | 258 (61.0) | 40 (48.2) | 115 (54.8) |
| I'm not sure | 53 (4.6) | 13 (3.0) | 20 (4.7) | 3 (3.6) | 17 (8.1) |
| **Awareness** |  |  |  |  |  |
| Q9. Awareness of leading cause of maternal mortality |  |  |  |  |  |
| Cancer | 13 (1.1) | 4 (0.9) | 4 (0.9) | 1 (1.2) | 4 (1.9) |
| Cardiovascular disease | 428 (37.1) | 160 (36.8) | 163 (38.5) | 29 (34.9) | 76 (36.2) |
| Infections | 36 (3.1) | 12 (2.8) | 13 (3.1) | 4 (4.8) | 7 (3.3) |
| Lack of access to quality healthcare | 484 (42.1) | 175 (40.2) | 185 (43.7) | 38 (45.8) | 86 (41.0) |
| Not sure | 171 (14.9) | 80 (18.4) | 49 (11.6) | 10 (12.0) | 32 (15.2) |
| Other | 19 (1.7) | 4 (0.9) | 9 (2.1) | 1 (1.2) | 5 (2.4) |

#### Awareness of Leading Cause of maternal Mortality among Respondents of Reproductive Age

Nearly half of participants (42.1%) identified lack of access to quality healthcare as the leading cause of maternal mortality **(Table 4)**. Over one-third of the respondents (37.1%) were aware that cardiovascular disease was the leading cause of maternal mortality, whilst a few attributed it to infections (3.1%) and cancer (1.1%).

### Postpartum Health

Among 423 postpartum participants, 82.0% recalled being screened for PPD. Of the 423 women, 13.0% reported a diagnosis of PPD, out of which 90.9% received treatment. There were no significant differences in PPD screening among different racial and ethnic groups: non-Hispanic White (82.9%), non-Hispanic Black (70.0%), Asian (75.0%), Hispanic (77.7%) **(Figure 2A)**. Similarly, there were no significant differences in PPD screening by educational status **(Figure 2B**) or income **(Figure 2C)**. Women who lived in a big city with access to many hospitals within 5 miles of home, had higher proportion of screening (88.7%), while rural area dwellers had the least (73.9%, *P =* 0.04) (**Figure 2D)**.

**Figure 2.**
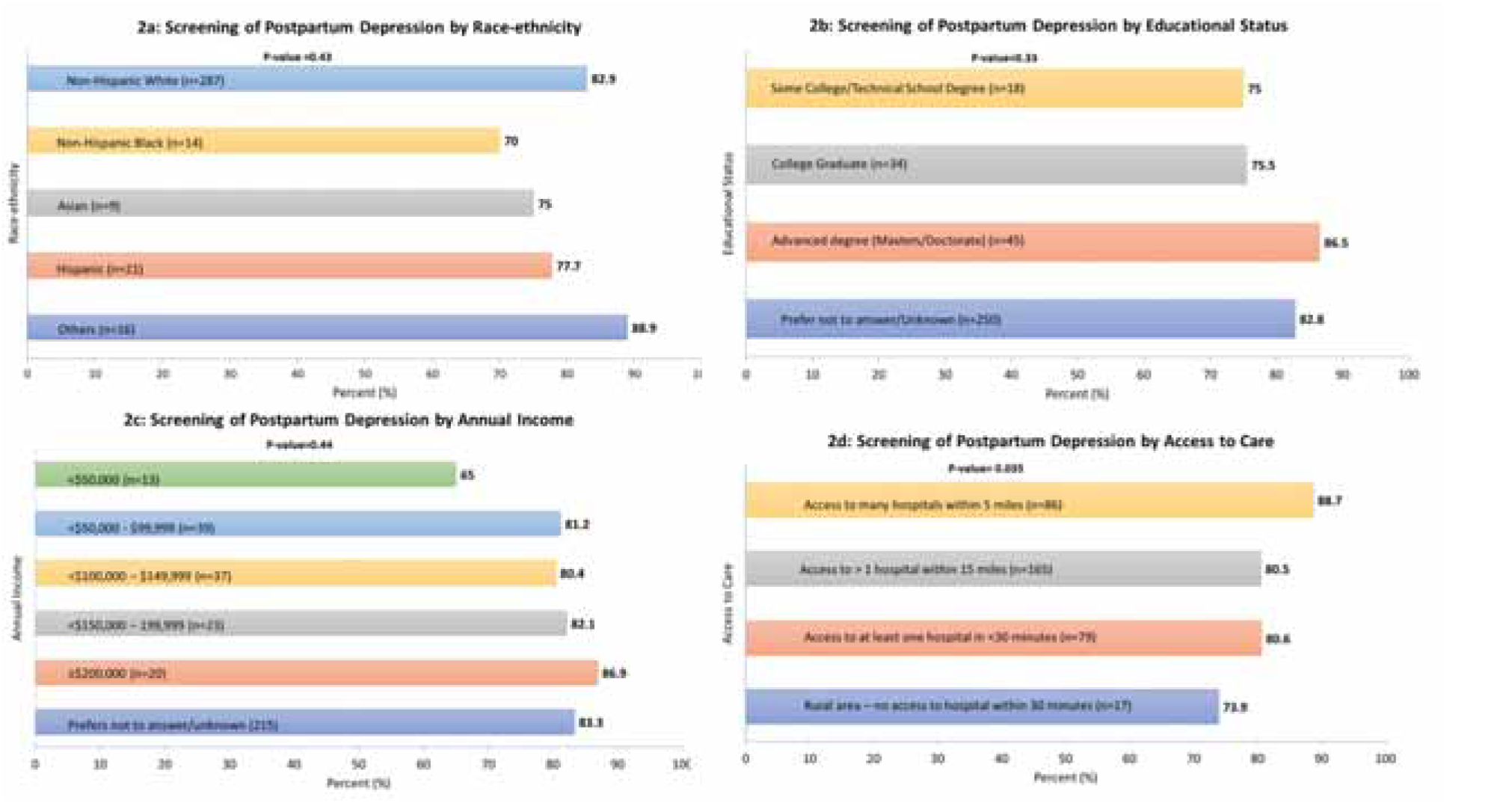
Screening for Pospartum depression by Race/ethnicity, educational status, income and access to care

## Discussion

To the best of our knowledge, this is the first cross-sectional study that evaluates pregnancy experiences, reproductive, and postpartum health and CVD awareness among a large registry across the US. This survey highlights the persistent gaps in reproductive care, education, and knowledge gaps among both patient and health care professionals. Nationally, 37% of participants were unaware of the association of APOs with long-term CVD risk, with significant variations by race. Additionally, 59% of participants reported that they were not educated on this association by their health care professionals. Pregnancy history and postpartum health screening gaps were also identified. Furthermore, 30% percent of postmenopausal individuals reported that their health care professionals did not take pregnancy history during recent visits, and this proportion varied significantly by race-ethnicity, income, and access to care, suggesting that socioeconomic status might be influencing the quality of prenatal care received. Only 37% of all individuals of reproductive age were aware that CVD was a leading cause of maternal mortality. Our key findings identified an urgent need for more education of the relationship between APOs and CVD to improve the reproductive experiences and CV awareness of individuals nationally.

Recent guidelines from the AHA, European Society of Cardiology, and American College of Obstetricians and Gynecologists, recommend the inclusion of APOs to evaluate long-term CVD risk in women.^12–16^ The guidelines recommend lifestyle modification and long-term follow-up, but do not state how APOs should be incorporated in risk assessment for CVD risk, likely because APOs are not included in any established risk scores for such risk assessment. Our data from respondents of reproductive age revealed that the most experienced adverse obstetric outcome was miscarriage, and majority of individuals did not receive preconception counseling prior to becoming pregnant. Our study found that 37% of individuals do not realize APOs are associated with long-term CVD risk, which is higher than what was reported in a recent cross-sectional study of over 5600 individuals, where 25% were unaware of an association between hypertensive disorders of pregnancy (gestational hypertension, preeclampsia, eclampsia) and CVD, and 24% of participants were unaware of an association between gestational diabetes mellitus and CVD.^17^ Furthermore, only 37% of individuals of reproductive age had awareness of CVD as a leading cause of maternal mortality, lower than what was reported in the same cross-sectional study, where 62% of respondents identified CVD as the leading cause of death in women.^17^ These discrepancies may be related to multiple factors such as respondent bias, recruitment site (academic medical center versus national survey), and varying level of education of respondents.^17^ Nevertheless, based on findings of both studies, at least 4 out of 10 participants were not able to correctly identify CVD as the leading cause of death in women, indicating that educational efforts in this direction are necessary and should be continued.

Further analysis of our data suggests that lack of education on this association from health care professionals is a major factor leading to this CVD risk unawareness. Thirty-five percent of postmenopausal individuals reported that their health care providers did not take pregnancy history during visits, and this proportion varied significantly by income and by race/ethnicity. According to a recent physician survey, only half of internal medicine and family medicine providers who responded, acknowledged awareness of the association between APOs and long-term CVD risk, compared to the vast majority of providers in OBGYN and cardiology acknowledging awareness of this association.^8^ The majority of providers not only fail to ask about APOs, but also lack the knowledge of how often to appropriately screen women for CVD risk factors that can be associated with APOs.^8^ A recent national survey of cardiologists, cardiovascular team members and cardiology fellows in training, identified that over 75% of respondents lacked access to a dedicated cardio-obstetrics team, and only 29% of cardiologists received didactics on cardio-obstetrics during training, suggesting that augmentation of cardio-obstetrics education is needed to reduce the large knowledge gaps that exist among clinicians.^18^ These data also identify a need for more education, both for physicians and for patients, on recognizing the association between APOs and long-term CVD risk in mothers.^6^

When analyzing data from a multiracial cohort of postpartum individuals, the prevalence of self-reported screening and diagnosis of PPD was 82% and 13%, respectively. Within the US, the prevalence of screening did not significantly differ by race, ethnicity, education, income, and proximity to health care. A recent study examining data from the 2009 – 2011 Pregnancy Risk Assessment Monitoring System examined racial/ethnic disparities in the relationship between antenatal stressful life events and PPD among women in the US (n = 87,565) and found a similar prevalence of diagnosis of PPD at 11%.^19^ Additionally, the authors found a protective effect of provider communication on PPD among non-Hispanic white and non-Hispanic black participants.^19^ Another survey of mothers in the Midwestern US (n = 61), examining the relationship between social support and PPD, has suggested that screening for depression alone may not be sufficient, as individuals may screen negative for PPD, but still report feelings of depression.^20^ In addition to using depression screening tools, a mother’s feelings should also be carefully assessed. This necessitates assessment of a mother’s physical and psychological health needs, as well as the nature of her support system.^20^ As such, it is imperative to educate healthcare professionals in the recognition and care of PPD using a multifaceted diagnostic approach. The prevalence of diagnosed PPD in the US did not significantly differ from the pooled global prevalence of diagnosed PPD, which based on recent studies is reported to be between 12% and 19%.^21–23^ Of these, a large systematic review and meta-analysis (n = 133,313), calculated a pooled prevalence of diagnosed PPD at 14% (95% confidence interval: 12 – 15%).^23^ The authors noted that the prevalence of diagnosed PPD varied according to country, with developing countries having a higher prevalence. The main risk factors associated with PPD were gestational diabetes mellitus, depression during pregnancy, male birth, history of depression during pregnancy, history of depression, and epidural anesthesia during delivery.^23^ These data offer healthcare professionals a theoretical basis for managing and treating patients with PPD, to help improve rates of screening, intervention and referral for individuals with PPD.

Our study had several strengths including data from a national registry of the AHA in the primary analytic sample, allowing us to reach a broad number of participants of childbearing age, as well as postpartum and postmenopausal individuals. This allowed us to effectively examine associations among respondents with high statistical power given large sample size. The sample was also recruited from individuals of the general public across the US, increasing the generalizability of the study to the overall national US population. However, the study also had some limitations. First, we utilized a survey that relied primarily on self-reporting of medical history, obstetric history, APOs, and personal experiences, introducing the potential for recall bias into the study design. design. Second, the data and conclusions drawn from therein are limited to respondents from the US and may not be generalizable on a global scale. Third, the online delivery of the survey platform may also have limited participation, due to a potential barrier from lack of internet access based on socioeconomic status of individuals across the nation as well as comfort level of individuals utilizing electronic platforms on digital devices.

In conclusion, considerable knowledge gaps exist on the association of APOs with CV risk, with disparities by race/ethnicity. Furthermore, most individuals are not educated on this association by their healthcare professionals. Hence, there is an urgent need for educating both patients and healthcare providers on the relationship between APOs and future CVD, to improve the health-care experiences and postpartum health outcomes of pregnant individuals.

## Data Availability

Data used in this manuscript can be made available upon reasonable request

## ABBREVIATIONS

APO: adverse pregnancy outcome
AHA-RGR: American Heart Association Research Goes Red
CVD: cardiovascular disease
PPD: postpartum depression

## Figure Legends

**Central Illustration.**
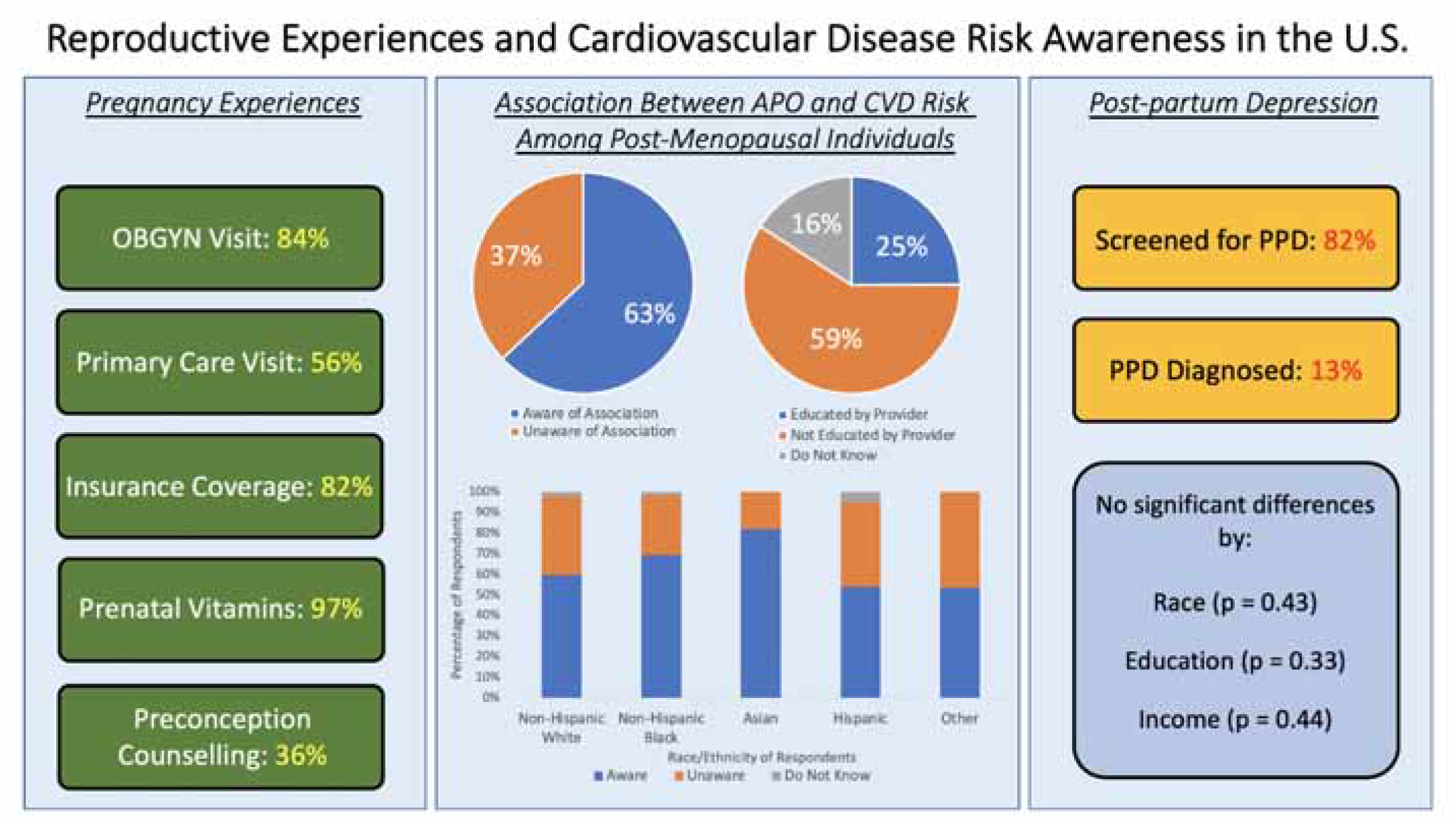
Reproductive Experiences and Cardiovascular Disease Risk Awareness in the U.S.

